# A Multicenter Evaluation of Blood Purification with Seraph 100 Microbind Affinity Blood Filter for the Treatment of Severe COVID-19: A Preliminary Report

**DOI:** 10.1101/2021.04.20.21255810

**Authors:** Stephen A. Chitty, Sarah Mobbs, Brian S. Rifkin, Steven W. Stogner, Michael S. Lewis, Jaime Betancourt, Jeffrey DellaVolpe, Fadi Abouzahr, Andrew M. Wilhelm, Harold M. Szerlip, Robert M. Gaeta, Ian Rivera, James D. Oliver, Stephen W. Olson, Subrata Debnath, Sean P. Barnett, Amay Parikh, Robert J. Walter, Mai T. Nguyen, Breandan Sullivan, Karl C. Alcover, Ian J. Stewart, Kathleen P. Pratt, Kumar Sharma, Kevin K. Chung, for the PURIFY INVESTIGATORS

**Affiliations:** Southeast Georgia Health System, Brunswick, GA; Hattiesburg Clinic, Hattiesburg, MS; Veterans Affairs Greater Los Angeles Healthcare System, Los Angeles, CA; Methodist Hospital, San Antonio, TX; University of Mississippi Medical Center, Jackson, MS; Baylor Scott & White Health, Dallas, TX; Dwight D. Eisenhower Army Medical Center, Fort Gordon, GA; Walter Reed National Military Medical Center, Bethesda, MD; University of Texas Health San Antonio, San Antonio, TX; Advent Health, Orlando, FL; Brooke Army Medical Center, San Antonio, TX; University of Colorado Anschutz, Aurora, CO; Uniformed Services University of the Heath Sciences, Bethesda, MD

**Keywords:** COVID-19, SARS-CoV-2, Hemoperfusion, Extracorporeal Circulation, Viremia, Critical Care Outcomes, Medical Countermeasure

## Abstract

**Objective:** The Seraph^®^100 Microbind Affinity Blood Filter^®^ (Seraph 100) is an extracorporeal medical countermeasure that can remove many pathogens from blood, including the SARS-CoV-2 virus. The aim of this study was to evaluate safety and efficacy of Seraph 100 treatment for severe coronavirus disease 2019 (COVID-19).

**Design:** Multicenter retrospective observational cohort study.

**Setting:** Intensive care units across four of thirteen participating sites who have completed data extraction.

**Patients:** Critically ill COVID-19 patients treated with Seraph 100 under an Emergency Use Authorization (n=53) and historical control patients who met criteria for treatment (n=46).

**Intervention:** Extracorporeal treatment with the Seraph 100 filter.

**Measurements and Main Results:** At baseline, the median age was 61 years, 72.7% were male, and 59.6% required mechanical ventilation. The groups were matched in terms of sex, race/ethnicity, body mass index, APACHE II score, need for mechanical ventilation, and other COVID-19 treatments. However, patients in the Seraph 100 group were younger with a median age of 61 years (IQR 42-65) compared to controls who had a median age of 64 (IQR 56-68, p=0.036). The Seraph 100 group also had a lower median Charlson comorbidity index (2, IQR 0-3) compared to control patients (3, IQR 2-4, p=0.006). Mortality was lower in the Seraph 100 treated group compared to the historical controls (37.7% vs 67.4%, respectively, p=0.003). Multivariable logistic regression analysis yielded an odds ratio of 0.27 (95% confidence interval 0.09-0.79, p=0.016). Of the 53 patients treated with Seraph 100, only 1 patient experienced a serious adverse event (transient hypotension at the start of the treatment which required a brief period of vasopressor support).

**Conclusions:** These data suggest that broad spectrum, pathogen agnostic, extracorporeal blood purification technologies can be safely and effectively deployed to meet new pathogen threats as an adjunct to standard treatments while awaiting the development of directed pharmacologic therapies and/or vaccines.

## Introduction

Coronavirus disease 2019 (COVID-19), caused by severe acute respiratory syndrome coronavirus-2 (SARS-CoV-2), is characterized by a profoundly dysregulated inflammatory response and concomitant endothelial dysfunction that results in end-organ damage.^1^ To date, SARS-CoV-2 has infected over 135 million people and killed almost 3 million world-wide.^2^ While advancements have been made in treating COVID-19, novel anti-viral therapeutics are still needed, particularly in those with critical illness.

For patients with sepsis, the development of ‘pathogenemia’ (i.e. bacteremia, viremia, fungemia) is consistently associated with worse outcomes.^3–6^ COVID-19 is no exception; emerging evidence suggests that SARS-CoV-2 viremia is common and directly linked to disease severity and poor outcomes. A recently published meta-analysis examined the association of viremia with outcomes, including data from 2,181 patients in 21 studies.^7^ The authors estimated that viremia occurs in 34% of patients and found that it was associated with COVID-19 severity. Furthermore, viremia was also associated with the risk of intensive care unit (ICU) admission, need for mechanical ventilation, multi-organ failure, and death. The strength of these associations was compelling, with odds ratios (OR) ranging from 4.3 for ICU admission to 11.1 for mortality. While causality cannot be determined from retrospective data, these results suggest that viremia itself may directly contribute to worse outcomes by allowing broad metastasis of viral invasion into non-pulmonary organs. Decreasing viremia in critically ill patients with COVID-19 might therefore improve outcomes.

The Seraph^®^100 Microbind Affinity Blood Filter^®^ (Seraph 100) (ExThera Medical, Martinez, CA) is an extracorporeal medical countermeasure designed to remove a multitude of pathogens from the blood. The Seraph 100 is a sorbent hemoperfusion filter containing polyethylene beads coated with immobilized heparin.^8^ This heparin surface mimics the endothelial glycocalyx and also allows for broad spectrum extracorporeal pathogen removal that is inclusive of viruses, bacteria, and fungi (see Supplemental Table 1). A recent report suggests that Seraph 100 is capable of removing SARS-CoV-2.^9^ Given prior work demonstrating the association between viremia and poor outcomes, clearance of SARS-CoV-2 from the bloodstream could be beneficial in critically ill patients with COVID-19 by providing adjunctive source control.

As a result of early experience with this device in patients with COVID-19^9^ accompanied by sufficient safety data, the Food and Drug Administration granted Emergency Use Authorization (EUA) for patients with COVID-19 with respiratory failure on 17 April 2020. We sought to collect data on patients treated under the EUA for a retrospective observational study to evaluate early evidence for safety and efficacy. We hypothesized that the treatment would be safe and associated with improved outcomes compared to historical controls. While enrollment is still ongoing, herein we present a preliminary analysis on the first group of study patients with complete data.

## Materials and Methods

The Blood purification with Seraph^®^100 Microbind Affinity Blood Filter^®^ for the treatment of severe COVID-19: An Observational Study (PURIFY-OBS-1) was reviewed and approved by the Advarra institutional review board in accordance with all applicable Federal regulations governing human research protections (Clinicaltirals.gov Identifier NCT04606498). The complete PURIFY-OBS-1 study includes three groups: 1) historical control group, 2) historical Seraph 100 treated patients, and 3) prospective Seraph 100 treated patients. To be included in the analysis in any of the groups, patients must have met the EUA criteria for treatment. These criteria required that patients be at least 18 years of age and have either: 1) early acute lung injury or acute respiratory distress syndrome, 2) severe disease (defined by dyspnea, respiratory rate >30 breaths/min, oxygen saturation ≤ 93%, or lung infiltrates >50%), or 3) life-threatening disease (respiratory failure, septic shock, or multi-organ dysfunction).

The historical control group was composed of patients that were admitted to the ICU at a participating site with COVID-19 that met inclusion criteria per the EUA but were not treated with the Seraph 100 device from 17 April 2020 (the date of EUA approval) until the protocol was approved at the study site. Since each site had slightly different clinical criteria for when they considered therapy with the Seraph 100 device, the investigators at each site were asked to identify all patients during the time period that they would have treated with the Seraph 100 device had it been available. The historical Seraph 100 treated cohort was composed of patient that were admitted to the ICU at a participating institution, had severe COVID-19 meeting EUA inclusion criteria, treatment duration of at least 4 hours up to 24 hours, and were admitted from the date of EUA approval (17 Apr 2020) until the date the protocol was approved at the study site. The third group, prospectively enrolled Seraph 100 treated patients is currently ongoing. This preliminary report only considers historical Seraph 100 and historical control patients treated at four centers in the United States. Exclusion criteria were: age greater than 75 years, incomplete survival data, and ICU admission greater than 7 days after hospital admission.

The sites collected data on admission to the ICU which included demographic variables (age, sex, race/ethnicity), body mass index (BMI), comorbid conditions (defined by the Charlson comorbidity index^10^ derived from chart review), and the acute physiology and chronic health evaluation (APACHE II) score.^11^ Data on other COVID-19 treatments, to include remdesivir and corticosteroids, were collected throughout the hospital stay. To assess outcomes, data on mortality, ICU length of stay, need for renal replacement therapy (RRT) on hospital discharge, and hospital length of stay were recorded. We also recorded laboratory data to assess blood indices and inflammatory markers. Blood indices of interest included white blood cell count (WBC), hemoglobin, and platelets. Inflammatory markers examined were C-reactive protein (CRP), ferritin, and D-dimer. Study personnel entered data into an on-line electronic data capture form in REDCap (Vanderbilt University, Nashville, TN).

Demographic distributions were compared using Chi-square test for categorical variables, unless cell sizes were small, in which case a Fisher’s exact test was utilized. For continuous variables, data were analyzed to determine distribution and none were normally distributed. Therefore, Mann-Whitney U tests were used to compare continuous variables between groups. The comparison of mortality rates between treatment and control groups was conducted using multiple logistic regression. Both univariate and multivariable analyses were performed to account for the potential confounding effects of age, sex, race/ethnicity, BMI, APACHE II score, and Charlson comorbidity index. In addition, comparison of survival rates was conducted and displayed graphically using a Kaplan-Meier survival curve and compared by means of a log-rank test. For the purposes of this analysis, if a patient was discharged alive before day 28, they were assumed to be alive at 28 days. To investigate the association between the length of treatment and differences in levels of blood indices and inflammatory markers, random effects models were used. The multilevel models treated each patient as the random variable. Associations were estimated using linear mixed effects regression models, with unstructured covariance matrix and time indicator as a covariate. Data were analyzed using Stata version 16.1 (StataCorp, College Station, TX).

## Results

In the 12 month period since April 2020, data were collected for a total of 61 patients who were admitted to the ICU with COVID-19 and treated with Seraph 100 across the four participating clinical sites. Data on an additional 84 patients were entered into the database to serve as historical controls for a total of N=145. Of these 145 subjects, 26 were excluded for missing data on mortality. An additional 17 patients were excluded for age >75 and 3 were excluded for ICU admission more than 7 days after hospital admission. The final study cohort was 99 subjects (N=53 for Seraph 100 treated and N=46 for controls). See Figure 1 for CONSORT diagram and Supplemental File 2 for study enrollment to date across all the participating sites.

**Figure 1.**
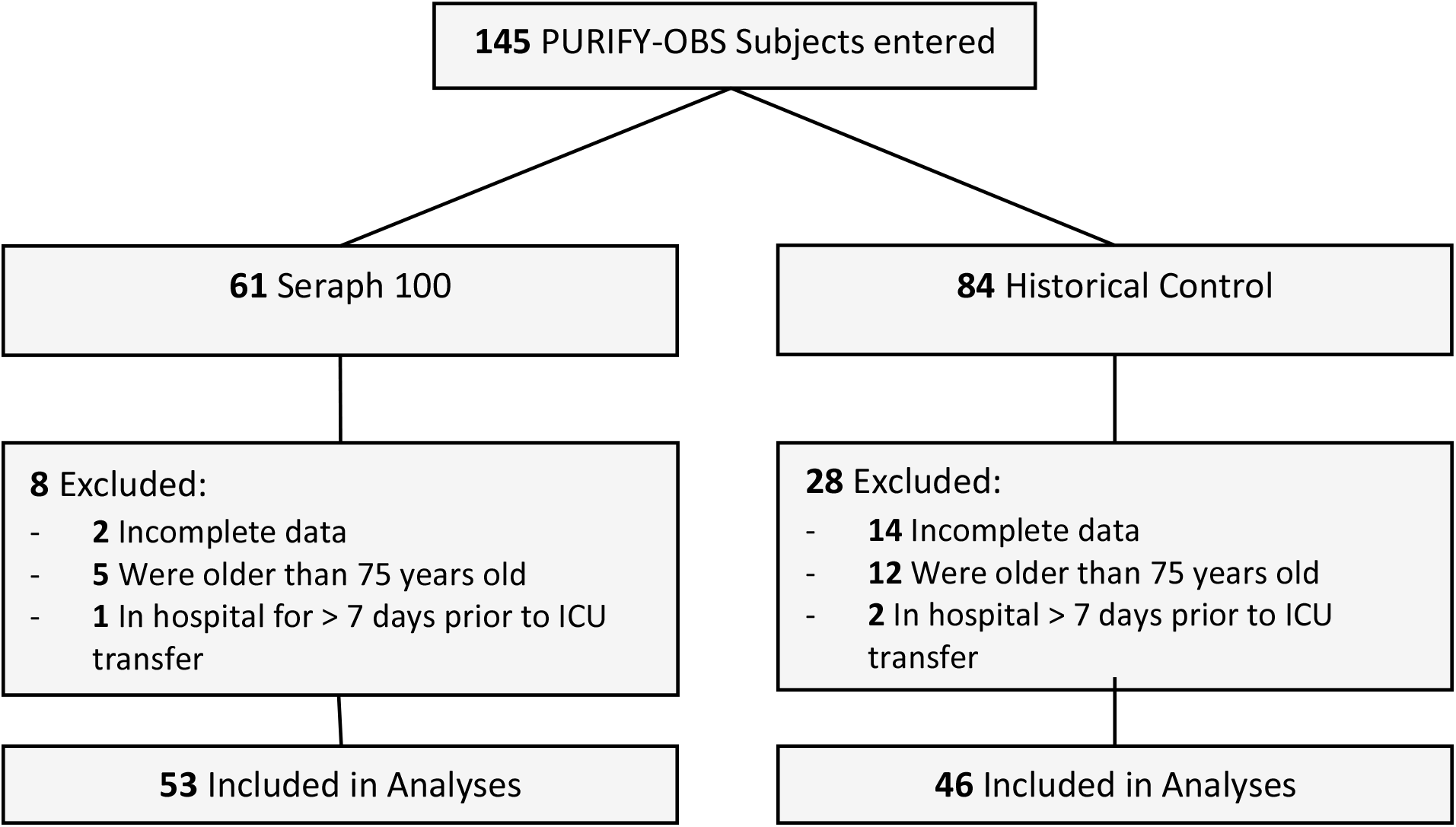
CONSORT Diagram

The baseline patient characteristics are presented in Table 1. Patients treated with Seraph 100 had a median age (Interquartile Range (IQR)) of 61 [42-65], which was significantly younger than controls 64 [56-68], p=0.036). The median Charlson comorbidity index (IQR) was higher in the control group (3 [2-4]) compared to the Seraph 100 treated group (2 [0-3], p=0.006). There was no significant difference noted between groups in terms of sex, race/ethnicity, BMI, APACHE II scores, or pre-existing diabetes. Similar numbers of subjects in both groups required mechanical ventilation (56.6% and 63.0% in Seraph 100 treated and control patients, respectively). The majority of patients in both groups were treated with remdesivir and corticosteroids. Additional data on available laboratory values at ICU admission are shown in Supplemental File 1, Table 2.

**Table 1.** Baseline patient characteristics

|  | Combined<br>(N=99) | Treatment<br>(N=53) | Control<br>(N=46) | P value |
| --- | --- | --- | --- | --- |
| Age, Median (IQR) | 61 (48-68) | 61 (42-65) | 64 (56-68) | 0.036 |
| Sex (%) |  |  |  | 0.510 |
| Male | 72.7 | 75.5 | 69.6 |  |
| Female | 27.3 | 24.5 | 30.4 |  |
| Race/Ethnicity (%) |  |  |  | 0.272 |
| NH White | 55.6 | 56.6 | 54.4 |  |
| NH Black | 19.2 | 22.6 | 15.2 |  |
| NH Asian/PI | 6.1 | 7.6 | 4.4 |  |
| Other | 4.0 | 5.7 | 2.2 |  |
| Unknown | 6.1 | 1.9 | 10.9 |  |
| Hispanic | 9.1 | 5.7 | 13.0 |  |
| BMI, Median (IQR)<sup>a</sup> | 34.0 (29.5-40.6) | 34.3 (29.7-40.7) | 33.3 (29.5-40.1) | 0.643 |
| APACHE II, Median (IQR) | 14 (10-19) | 13 (9-18) | 15.5 (11-21) | 0.059 |
| Charlson Comorbidity Index,<br>Median (IQR)<sup>b</sup> | 3 (1-4) | 2 (0-3) | 3 (2-4) | 0.006 |
| Diabetes (%)<sup>b</sup> | 39.8 | 34.6 | 45.7 | 0.265 |
| Mechanical<br>Ventilation (%)<sup>c</sup> | 59.6 | 56.6 | 63.0 | 0.515 |
| COVID Treatments |  |  |  |  |
| Remdesivir (%) | 77.8 | 83.0 | 71.1 | 0.178 |
| Corticosteroids (%) | 97.0 | 100.0 | 93.5 | 0.097 |

NH= Non-Hispanic
PI= Pacific Islander
<sup>a</sup>Data missing for two study subjects
<sup>b</sup>Data missing for one study subject
<sup>c</sup>At anytime within the first 7 days of intensive care unit admission

**Table 2.** Outcomes stratified by study cohort

|  | Treatment | Control | P value |
| --- | --- | --- | --- |
| Mortality (%) | 37.7 | 67.4 | 0.003 |
| ICU-free days, Median (IQR)<sup>a</sup> | 10.5 (0-19.5) | 0 (0-12.5) | 0.052 |
| RRT dependent at discharge (%)<sup>b</sup> | 0 | 9.4 | 0.541 |
| Hospital length of stay (day), Median (IQR)<sup>b</sup> | 17 (10-35.5) | 15 (5-32) | 0.170 |

RRT= renal replacement therapy
<sup>a</sup>Data available for 88 subjects
<sup>b</sup>Among survivors, data missing for one study subject

Outcomes of interest are presented in Table 2. More subjects in the control group died (67.4%) compared to patients treated with Seraph 100 (37.3%, p=0.003). A Kaplan-Meier curve stratified by group demonstrating survival over time is presented in Figure 2 (p<0.001). There were no significant differences noted for 28 day-ICU free survival, RRT dependence on discharge, or hospital length of stay.

**Figure 2.**
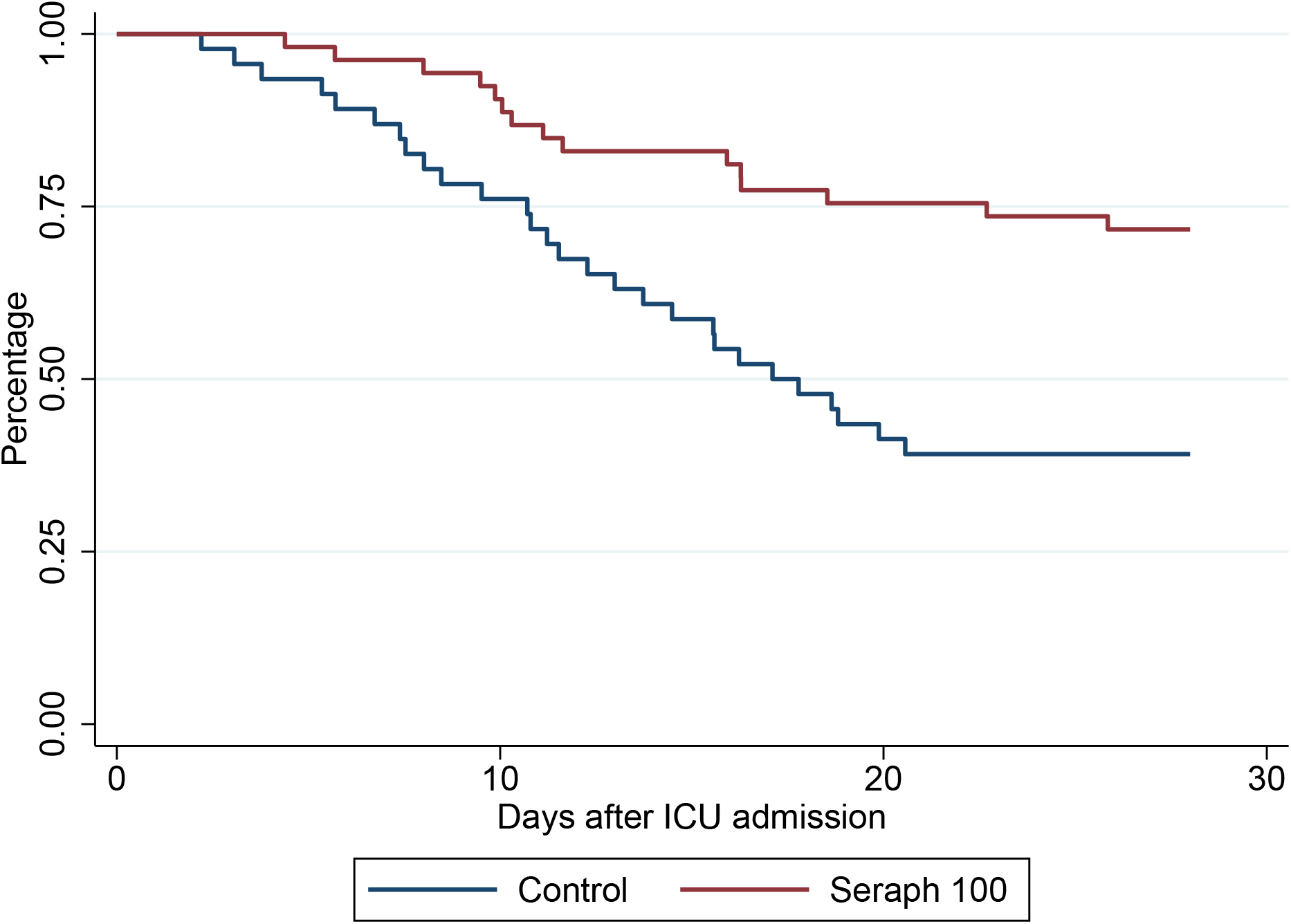
Kaplan-Meier survival curve stratified by treatment

The results from the univariate and multivariable logistic regression models are shown in Table 3. On univariate analysis, treatment with Seraph 100 was associated with a decrease in mortality with an OR of 0.29 and 95% confidence interval (CI) of 0.13-0.67; p=0.004. Other significant variables in the univariate analysis were age (OR 1.09 per one year increase, 95% CI 1.05-1.14; p<0.001), APACHE II (OR 1.11 per one point increase, 95% CI 1.04-1.19; p=0.001), and Charlson comorbidity index (OR 1.47 per one point increase, 95% CI 1.17-1.84; p=0.001). Sex, race/ethnicity, and BMI were not associated with mortality in the univariate analysis. In the multivariable model, treatment with the Seraph 100 device (OR 0.27, 95% CI 0.09-0.79; p=0.016) and age (OR 1.09, 95% CI 1.01-1.17; p=0.024) remained significant. APACHE II and Charlson comorbidity index were no longer significant after adjustment.

**Table 3.** Univariate and multivariable models for the outcome of mortality

|  | Univariate |  |  | Multivariable<sup>a</sup> |  |  |
| --- | --- | --- | --- | --- | --- | --- |
|  | Odds Ratio | 95% CI | P value | Odds Ratio | 95% CI | P value |
| Treatment |  |  |  |  |  |  |
| Control | Ref | - | - | Ref | - | - |
| Seraph 100<sup>b</sup> | 0.29 | 0.13-0.67 | 0.004 | 0.27 | 0.09-0.79 | 0.016 |
| Age (per year) | 1.09 | 1.05-1.14 | <0.001 | 1.09 | 1.01-1.17 | 0.024 |
| Sex |  |  |  |  |  |  |
| Female | Ref | - | - | Ref | - | - |
| Male | 0.42 | 0.17-1.07 | 0.068 | 0.66 | 0.21-2.09 | 0.478 |
| Race/Ethnicity |  |  |  |  |  |  |
| NH White | Ref | - | - | Ref | - | - |
| NH Black | 0.93 | 0.33-2.63 | 0.885 | 1.36 | 0.37-5.05 | 0.647 |
| NH Asian/PI | 0.83 | 0.15-4.50 | 0.832 | 1.02 | 0.15-7.15 | 0.981 |
| Other | 0.83 | 0.11-6.35 | 0.860 | 1.41 | 0.13-15.38 | 0.778 |
| Unknown | 1.67 | 0.28-9.87 | 0.573 | 0.89 | 0.09-8.61 | 0.922 |
| Hispanic | 0.24 | 0.05-1.25 | 0.090 | 0.29 | 0.04-2.26 | 0.240 |
| BMI (per 1 point increase) | 0.99 | 0.94-1.03 | 0.562 | 1.02 | 0.95-1.09 | 0.566 |
| APACHE II (per 1 point increase) | 1.11 | 1.04-1.19 | 0.003 | 1.03 | 0.95-1.09 | 0.466 |
| Charlson Comorbidity Index | 1.47 | 1.17-1.84 | 0.001 | 0.96 | 0.68-1.36 | 0.832 |

<sup>a</sup>N for multivariable model is 97
<sup>b</sup>Compared to historical control

The linear mixed effects regression models for blood indices and inflammatory markers are presented in Table 4. These models demonstrated that treatment time was not significantly associated with changes in WBCs or hemoglobin, but longer times were associated with an increase in platelets compared to shorter treatments (p=0.003). Treatment time was not associated with changes in CRP, ferritin, or D-dimer.

**Table 4.** Associations between length of treatments and changes in blood indices and inflammatory markers

|  | Coefficient | 95% Confidence Interval | P value |
| --- | --- | --- | --- |
| Blood Indices<sup>a</sup> |  |  |  |
| White Blood Cells | 2.15 | -0.81-5.10 | 0.154 |
| Hemoglobin | 0.06 | -0.62-0.74 | 0.864 |
| Platelets | 52.79 | 18.56-87.01 | 0.003 |
| Inflammatory Markers |  |  |  |
| C-Reactive Protein<sup>b</sup> | 0.86 | -4.32-6.05 | 0.744 |
| Ferritin<sup>c</sup> | 147.40 | -216.88-511.69 | 0.428 |
| D-dimer<sup>d</sup> | -0.42 | -2.60-1.75 | 0.707 |

<sup>a</sup>Data from 67 treatments in 30 patients
<sup>b</sup>Data from 64 treatments in 29 patients
<sup>c</sup>Data from 46 treatments in 25 patients
<sup>d</sup>Data from 67 treatments in 33 patients

With respect to our safety outcome, only one adverse event occurred during treatment with Seraph 100. This was an episode of hypotension that required initiation of norepinephrine. The patient subsequently survived and was discharged from the hospital nine days after the episode.

## Discussion

In this initial report of the Seraph 100 for the treatment of severe COVID-19, we observed that treatment with the Seraph 100 device decreased mortality compared to contemporaneous controls. This survival benefit remained statistically significant after adjustment for demographics, BMI, APACHE II, and Charlson comorbidity index. Furthermore, we found that treatment with the device was safe, associated with only one serious adverse event. We did not observe differences in 28 day ICU-free mortality, need to RRT on discharge, or hospital length of stay. Length of treatment did not appear to impact changes in inflammatory markers or blood indices, with the exception of platelets.

As the COVID-19 pandemic has evolved, a few therapeutic agents have been shown to have important impacts on outcomes. Remdesivir has been found to be beneficial when given early in the disease course, prior to patients requiring advanced respiratory support.^12^ Tocilizumab, a monoclonal antibody that binds to the interleukin-6 receptor, is another promising therapy that has been shown to decrease mortality in hospitalized patients among those requiring advanced respiratory support.^13^ Glucocorticoid therapy as demonstrated mortality benefit across illness severity among critically ill patients.^14,15^ The RECOVERY trial demonstrated a 17% decrease in the age-adjusted rate ratio for mortality in patients treated with dexamethasone.^15^ While we did not capture Tocilizumab data in our preliminary data pull, most of our patients received remdesivir (83% and 71.1% in the Seraph 100 group and control group, respectively) while nearly all patients received glucocorticoids (100% and 93.5% in the Seraph 100 and control group, respectively). Our preliminary results provide evidence that extracorporeal blood purification with Seraph 100 could be a useful adjunct to standard pharmacologic therapies to improve outcomes.

The notion that infection can be treated with an extracorporeal approach is novel as the foundation of the treatment of infection for seven decades has been antimicrobial therapy. However, the first tenet of sepsis treatment is source control.^16^ In patients with either new pathogens or pathogens with high levels of resistance, antimicrobials are ineffective. The concept of a dialysis-like therapeutic is to enhance source control is rational. For example, when a drain is placed into an abscess which removes large amounts of infected material, some purulent material remains for the immune system to clear. Similarly, the Seraph 100 is an adjunctive treatment to clear the bloodstream of pathogen. This concept of debulking or blood stream clearance has been utilized to treat malaria and babesiosis when the pathogen burden is high, even in a background of effective anti-microbial treatment.^17^ Thus, the Seraph 100 was developed as an extracorporeal medical countermeasure that can be utilized as adjunctive therapy for a multitude of pathogens (see Supplemental File 1, Table 1). SARS-CoV-2 requires heparin/heparan sulfate to bind to cells^18^, thus the Seraph 100 is likely to remain highly effective for SARS-CoV-2 blood clearance regardless of the COVID-19 variant. This point is important as COVID-19 variants have already demonstrated immune escape from vaccines and monoclonal antibodies.^19^ In addition, remdesivir resistance is also possible.^20^

Although our study demonstrates promising results, limitations exist. Firstly, this is a retrospective analysis and not a randomized controlled trial. Since each study site had slightly different local criteria for initiating therapy with Seraph 100, we were unable to establish standardized criteria for historical control subjects. Each site was asked to apply their clinical practice criteria to patients treated at their institution prior to the availability of the Seraph 100 at their site to select control patients. This introduces the possibility of selection bias in the control group. Secondly, treatment times were not standardized and ranged from 15 minutes to over 2 days. Lastly, while the majority of patients in both groups were treated with remdesivir and glucocorticoids, these treatments were not standardized and data on other treatments (such as convalescent plasma, baricitinib, and tocilizumab) were not recorded.

## Conclusions

In conclusion, we found that a non-pharmacologic medical countermeasure, Seraph 100, was safely deployed during the COVID-19 pandemic. This initial assessment with contemporaneous control patients showed statistically significant improvement survival in severely ill COVID-19 patients even after controlling for confounders. These data suggest that a broad spectrum, pathogen agnostic, extracorporeal, blood purification device can be safely and effectively deployed to meet new pathogen threats as an adjunct to standard treatments while awaiting the development of directed pharmacologic countermeasures or vaccines. Complete data for PURIFY-OBS-1 will be forthcoming. Additionally, we will soon launch a multicenter, randomized controlled feasibility trial of the Seraph 100 for septic shock due to any pathogen.

## Supporting information

Supplemental File 2 - Enrollment

Supplemental File 3 - PURIFY INVESTIGATORS

## Data Availability

Raw data will be available once final report is published upon request.

## Acknowledgments

Please see the Supplemental File 3 for a full list of PURIFY-OBS-1 Investigators. The authors would like to thank the ICU physicians, ICU nurses, and dialysis nurses, without whom the extracorporeal treatments would not have been possible. We also thank the patients and their families, who put their trust in this experimental device. Lastly, we would like to thank the Defense Advanced Research Projects Agency, who provided early funding support for the Seraph 100 device.

