## Supplemental File 1 - Tables for "A Multicenter Evaluation of Blood Purification with Seraph 100 Microbind Affinity Blood Filter for the Treatment of Severe COVID-19: A Preliminary Report"

Table 1. Available laboratory values on admission to the intensive care unit

|  | **Treatment** | **Control** | **P value** |
| --- | --- | --- | --- |
| White Blood Cell Count (10^3^/uL)  N=55 | 9.5 (7.0-11.2) | 8.4 (6.1-12.1) | 0.543 |
| Hemoglobin (g/dL)  N=55 | 13.5±1.4 | 13.0±2.9 | 0.407 |
| Platelets (10^3^/uL)  N=55 | 250.5 (200-286) | 235 (177-284) | 0.600 |
| C-Reactive Protein (mg/L)  N=42 | 14.7 (4.8-23.6) | 14.7 (8.8-23.4) | 0.421 |
| Ferritin (mcg/L)  N=17 | 807.1 (352.0-1030.0) | 808.7 (257.9-1797.3) | 0.845 |
| D-Dimer (ng/mL)  N=43 | 1.2 (0.5-2.8) | 1.3 (0.8-2.5) | 0.451 |
| Sodium (mmol/L)  N=62 | 137 (134.5-138) | 135 (132-138) | 0.357 |
| Potassium (mmol/L)  N=62 | 4.1 (3.7-4.5) | 4.1 (3.8-4.7) | 0.616 |
| Chloride (mmol/L)  N=62 | 99.5 (96.5-103) | 99 (96-103) | 0.735 |
| Total CO_2_ (mmol/L)  N=62 | 27.2±4.8 | 24.7±5.5 | 0.064 |
| Blood Urea Nitrogen (mg/dL)  N=62 | 19.5 (14-25) | 20.5 (13-36) | 0.442 |
| Creatinine (mg/dL)  N=62 | 0.9 (0.7-1.3) | 1.0 (0.8-1.4) | 0.333 |
| Glucose (mg/dL)  N=62 | 137 (116-167) | 152 (123-217) | 0.418 |
| Calcium (mg/dL)  N=56 | 8.5±0.4 | 8.3±0.6 | 0.217 |
| pH  N=40 | 7.42 (7.39-7.46) | 7.36 (7.31-7.40) | 0.009 |
| pCO_2_ (mmHg)  N=40 | 38.5±11.5 | 46.8±17.2 | 0.076 |
| pO_2_ (mmHg)  N=40 | 67 (55-79) | 68 (63-82) | 0.692 |
| FiO2  N=88 | 1.0 (0.8-1.0) | 1.0 (0.8-1.0) | 0.590 |
| Aspartate Aminotransferase (U/L)  N=55 | 57 (43-87) | 66 (47.5-115.5) | 0.429 |
| Alanine Aminotransferase (U/L)  N=54 | 49 (22-82) | 40 (19-59) | 0.489 |
| Albumin (g/dL)  N=55 | 3.7±0.5 | 3.5±0.4 | 0.033 |
| Total Protein (g/dL)  N=50 | 7.3±0.8 | 6.8±0.6 | 0.017 |
| Total Bilirubin (mg/dL)  N=55 | 0.8 (0.6-1.1) | 0.6 (0.5-0.9) | 0.063 |
| Alkaline Phosphatase (U/L)  N=54 | 76 (55-98) | 72 (56-95) | 0.829 |
| Prothrombin Time (seconds)  N=17 | 14.6±1.5 | 14.6±1.6 | 0.949 |
| INR (seconds)  N=17 | 1.1±0.1 | 1.1±0.2 | 0.923 |
| Lactate (mmol/L)  N=20 | 1.4 (1.2-2.3) | 1.5 (1.0-1.8) | 0.705 |

Data reported as median (interquartile range) or mean ± standard deviation as appropriate for data distribution and compared by Mann-Whitney U tests and t-tests, respectively

FiO2: fraction of inspired oxygen

pCO_2_: partial pressure of carbon dioxide

pO_2_: partial pressure of oxygen

INR: International Normalized Ratio

Table 2. Pathogens removed by Seraph 100

| **Pathogens and heparin binding sepsis mediators** | **Single Pass Binding (%)** |
| --- | --- |
| **Bacteria^a^** |  |
| MRSA | 92 |
| *K. pneumoniae* (CRE) | 99.9 |
| *K. pneumoniae* | 37 |
| *E. coli* (CRE) | 99.9 |
| *E. coli* | 99.7 |
| *S. pneumoniae* | 53 |
| *E. faecalis* | 99.0 |
| *E. faecalis* (VRE) | 91 |
| *E. faecium* | 56 |
| *A. baumannii* | 79 |
| *S. epidermidis* | 58 |
| Methicilin resistant *S. epidermidis* | 66 |
| *S. pyogenes* | 76 |
| *Serratia marcescens* | 73 |
| **Viruses** |  |
| Ebola ^b^ | (39-99) |
| Zika ^a^ | 87 |
| Adenovirus ^a^ | 62 |
| Cytomegalovirus (CMV) ^a^ | 79 |
| **Fungi** |  |
| *C. albicans* ^b^ | 36 |

^a^Seffer, Malin-Theres, et al. "Heparin 2.0: a new approach to the infection crisis." Blood Purification 50.1 (2021): 28-34.

^b^ExThera Medical Internal Reports
