## Supplemental File 2 - Enrollment for "A Multicenter Evaluation of Blood Purification with Seraph 100 Microbind Affinity Blood Filter for the Treatment of Severe COVID-19: A Preliminary Report"

**PURIFY-OBS-1 Enrollment Update – (*denotes activated site)**

| **Rank** | **Site** | **Seraph 100 Treated under EUA**  **(Eligible for PURIFY-OBS)** | **Retrospective**  **Seraph 100** | **Historical**  **Control** | **Prospective**  **Seraph 100** | **TOTAL**  **ENROLLED** |
| --- | --- | --- | --- | --- | --- | --- |
| 1 | *Southeast Georgia (SE GA) | 60 | 34 | 32 | 0 | 66 |
| 2 | *Hattiesburg Clinic | 31 | 20 | 20 | 0 | 40 |
| 3 | *Los Angeles Veterans Affairs (LA VA) | 8 | 5 | 30 | 0 | 35 |
| 4 | *Methodist San Antonio | 6 | 2 | 2 | 0 | 4 |
| 5 | *University of Mississippi | 9 | 0 | 0 | 0 | 0 |
| 6 | *Baylor Scott & White | 4 |  |  |  |  |
| 7 | *University of Texas Southwestern | 0 |  |  |  |  |
| 8 | Eisenhower Army Medical Center (EAMC) | 21 |  |  |  |  |
| 9 | Walter Reed | 4 |  |  |  |  |
| 10 | University of Texas Health San Antonio (UTHSA) | 3 |  |  |  |  |
| 11 | Advent Health - Orlando | 3 |  |  |  |  |
| 12 | Brooke Army Medical Center | 1 |  |  |  |  |
| 13 | Colorado - Anschutz | 1 |  |  |  |  |
|  | **TOTAL** | **151** | **61** | **84** | **0** | **145** |
