## Supplemental File 3 - PURIFY INVESTIGATORS for "A Multicenter Evaluation of Blood Purification with Seraph 100 Microbind Affinity Blood Filter for the Treatment of Severe COVID-19: A Preliminary Report"

Amay Parikh, MD, MBA, MS

Sarah Guyler, BS, MHA, CCRP

**Baylor Scott & White Health, Dallas, TX**

Harold Szerlip, MD

Giselle Carino

Tanqunisha Coleman

**Brooke Army Medical Center, San Antonio, TX**

Robert J. Walter, MD, DHCE

Mai Nguyen, MD

**Dwight D. Eisenhower Army Medical Center, Fort Gordon, GA**

Robert Gaeta, DO

Ian M. Rivera, MD

Ferdinand Bacomo, DO

Susan D. Rogers, BS, CCRC

**ExThera**

Bob Ward, PhD

Keith McCrea, PhD

Erdie de Peralta, MA

Lakhmir S. Chawla, MD

Andrea Worsham, BS, MA

Michael Benjamin

**Hattiesburg Clinic, Hattiesburg, MS**

Brian Rifkin, MD

Steven Stogner, MD

Steve Farrell, MD

Karen Brooks, RN

**Henry Jackson Foundation**

Maria Voelkel

Sophie Haralson, RRT, MS, PMP

Marianne Spevak, BSHS, CCRC, CCRA

**HJF-ACESO**

Daniell Clark, PhD

Laura Osgood

Qianru Wu, MS

Kevin Grant

Rittal Mehta

**Methodist Hospital, San Antonio, TX**

Jeffrey Dellavolpe, MD

Fadi Abouzahr, MD

Mohammed Ahmed, MD

Ginger Dowell, ACNP

Stephen Amerson

**Southeast Georgia Health System, Brunswick, GA**

Stephen Chitty , MD

Srah Mobbs, FNP-BC

Gwen Gratto-Cox

Christy Jordan, RN, JD

**Trauma Insight**

George Peoples, MD, FACS

Dan Hargrove, JD, LLM

Karen Arrington, RN

Lauren Zahra, MS

Jessica Raley, PhD

Katie Lyon, MS, CCRP, CPM, CRMP

Rachel Macomber

Ashlee Richie, CCRP, CCDM

Tineka Brown

Laura Richie

Susan Hargrove, JD

Emily Scribe, PharmD

Steven White, JD

**Uniformed Services University of the Health Sciences, Bethesda, MD**

Kevin Chung, MD, FCCM, FACP

Ian Stewart, MD

Kathleen Pratt, PhD

Karl Alcover, PhD

Jason Lees, PhD

Amy Laczek

**University of Colorado Anschutz, Aurora, CO**

Breandan Sullivan, MD

Nick Naughton, MPH

Sabrina Espinoza

Vikhyat Bebarta, MD

Adit Ginde, MD, MPH

Anip Bansal, MD

Ilona Dewald, MPH

Isidro Susano Basilio

**University of Mississippi Medical Center, Jackson, MS**

Andrew Wilhelm, DO

Delia Owens, MSN, JD, RN, CCRP

**University of Texas Health San Antonio, San Antonio, TX**

Kumar Sharma, MD

Subrata Debnath, MBBS, MPH, PhD

Sean P. Barnett, MD

**University of Texas Southwestern, Dallas, TX**

Benjamin Levi, MD

Caroline Park, MD

Tamim Hamdi. MD

Peiman Lahsaei, MD

Nilum Rajora, MD

Christopher Choi, MD

**Veterans Affairs Greater Los Angeles Healthcare System, Los Angeles, CA**

Michael Lewis, MD

Jaime Betancourt, MD

Nancy Mohler

Natalia Dudek

Jasmine Bagnas

**Walter Reed National Military Medical Center, Bethesda, MD**

James Oliver, MD, PhD

Stephen Olson, MD

Jenny Nguyen
